# Genotype-guided isoniazid dosing harmonizes drug exposure in 3HP tuberculosis preventive therapy

**DOI:** 10.64898/2026.08.27.26360825

**Authors:** Kesia Esther da Silva, Samuel Sarkodie, Karina Marques, Patricia Vieira, Roberto Dias de Oliveira, Paulo César Pereira dos Santos, Marco Antonio Moreira Puga, Allyson Guimarães Costa, João Paulo Gregorio Machado, Renata Spener-Gomes, Eunsol Yang, Rada Savic, Marcelo Cordeiro-Santos, Julio Croda, Jason R. Andrews

**Author notes:** Correspondence: Jason Andrews, Division of Infectious Diseases and Geographic Medicine Stanford University School of Medicine Biomedical Innovations Building Stanford University School of Medicine Stanford, CA, 94305. Co-senior authors (Julio Croda, Jason R. Andrews). ClinicalTrials.gov: NCT05413551.

## Abstract

**Rationale:** Polymorphisms in the N-acetyltransferase 2 (NAT2) gene explain much of the interindividual variation in isoniazid (INH) metabolism and determine risk of toxicities. However, there is limited evidence to guide INH dose adjustment according to the NAT2 acetylator profile in weekly rifapentine-INH tuberculosis preventive therapy (TPT).

**Objectives:** We evaluated whether NAT2 genotype-guided INH dose adjustment harmonizes drug exposure across acetylator phenotypes during 3HP and derived phenotype-specific dose recommendations.

**Methods:** In a prospective, multicenter, within-subject PK trial (NCT05413551), adults initiating 3HP in Brazil were assigned genotype-guided INH doses (slow: 5 mg/kg ≤300 mg; intermediate: 15 mg/kg ≤900 mg; rapid: 25 mg/kg ≤1,500 mg) alongside a standard 900 mg flat dose on an alternate occasion. AUC0–24 and C24 were estimated from serial blood samples; a two-compartment Michaelis-Menten population PK model characterized NAT2 effects on clearance.

**Measurements and Main Results:** Among 228 participants, 47.4% (108/228) were intermediate, 43.4% (99/228) slow, and 9.2% (21/228) rapid acetylators. Genotype-guided dosing reduced AUC0–24 variability approximately two-fold versus standard dosing (CV 58.8% vs 76.8%) and increased exposure uniformity (median AUC0– 24 27.2 [IQR 18.8–41.3] vs 43.2 [27.3–71.0] mg·h/L). Among slow acetylators, C24 >0.15 µg/mL decreased from 27/42 (64%) with standard dosing to 1/42 (2%) with genotype-guided dosing (P<0.0001). In 104 participants with intensive PK sampling, rapid acetylators receiving guided doses had AUC0–24similar to standard-dose intermediate acetylators (42.8 vs 39.5 mg·h/L; P=.63). Monte Carlo simulations supported doses of 600, 900, and 1,200 mg for slow, intermediate, and rapid acetylators, respectively.

**Conclusions:** NAT2-guided isoniazid dosing reduced variation in drug levels, averting very low and high AUC and C_24_. These findings inform genotype-stratified dosing of INH for TPT, which might reduce toxicities and improve outcomes.

## Introduction

Tuberculosis (TB) remains the leading cause of death from a single infectious agent worldwide, causing an estimated 1.23 million deaths in 2024.[1] Twelve once-weekly doses of isoniazid (INH, 15 mg/kg) plus rifapentine (3HP) is the preferred short-course tuberculosis preventive therapy (TPT) owing to superior completion rates and non-inferior efficacy versus longer daily regimens.[4] However, in the largest programmatic cohort, 36% of participants experienced at least one adverse event,[5] and randomized trials have shown a nine-fold higher odds of systemic drug reactions (SDRs) with 3HP than with daily INH.[6] As 3HP is rolled out at scale, optimizing dosing to preserve efficacy while reducing toxicity is increasingly urgent.

INH is metabolized by arylamine N-acetyltransferase 2 (NAT2); polymorphisms in NAT2 explain more than 80% of interindividual INH pharmacokinetic variability, classifying individuals as slow, intermediate, or rapid acetylators.[7] Slow acetylators have a three-to four-fold increased risk of INH-induced liver injury[9, 10] and a seven-fold higher risk of SDRs during 3HP,[11] while rapid acetylators are prone to subtherapeutic exposure and increased microbiologic failure, and relapse.[12, 13] This evidence is restricted to daily INH dosing; no prior study has evaluated NAT2-guided dose modification for the higher weekly 3HP dose, which produces the peak concentrations that drive SDRs.[11]

In this study, we conducted a prospective *NAT2*-guided PK trial of weekly INH-rifapentine in a diverse Brazilian cohort initiating treatment for TB infection. Participants received phenotype-guided INH doses (slow: 5 mg/kg; intermediate: 15 mg/kg; rapid: 25 mg/kg) alongside a standard 900 mg reference occasion. Using population PK modeling, we compared INH exposure under genotype-guided versus standard flat dosing to evaluate whether NAT2-guided dosing harmonizes INH exposure across acetylator phenotypes.We evaluated whether NAT2-guided dosing harmonizes INH exposure across acetylator phenotypes.

## Methods

### Study design

This was a prospective, multicenter, open-label, pharmacogenomic-guided PK trial of the 3-month weekly INH plus rifapentine (3HP) regimen for the treatment of *M. tuberculosis* infection (TBI), conducted in Brazil (NCT05413551). The protocol was reviewed and approved by the Research Ethics Committees of the Federal University of Mato Grosso do Sul (UFMS) and the Dr. Heitor Vieira Dourado Tropical Medicine Foundation (FMT-HVD), the Brazilian National Committee for Ethics in Research (CAAE 58751622.8.0000.0021), and the Stanford University Institutional Review Board. The study followed the Declaration of Helsinki and ICH-GCP guidelines, and all participants provided written informed consent.

### Study sites and participants

Participants were enrolled across two cities in Brazil. In Campo Grande, Mato Grosso do Sul, enrollment occurred at UFMS clinics and two prison units; in Manaus, Amazonas, enrollment occurred at the FMT-HVD. Participants were recruited from five groups eligible for TPT under Brazilian guidelines: healthcare workers with TB infection (BRACE trial), prison staff, people deprived of liberty, household contacts of pulmonary TB cases, and people living with HIV (PLHIV). The first three groups were recruited at Campo Grande only; household contacts and PLHIV were enrolled at both sites.

Eligible participants were adults aged ≥18 years with evidence of TBI, defined as a positive QuantiFERON-TB Gold Plus interferon-γ release assay (IGRA) or, for PLHIV with CD4 <350 cells/mm³, eligibility for TPT irrespective of IGRA result. Screening included clinical evaluation, chest radiography, and, when indicated by symptoms or radiographic abnormalities, sputum Xpert MTB/RIF Ultra and mycobacterial culture to rule out active TB. Key exclusion criteria were evidence of active TB; prior treatment for active or TBI for more than 14 days; close contact with rifampicin-or INH-resistant TB; known intolerance or hypersensitivity to INH or rifapentine; clinical diagnosis of active liver disease or alcohol dependence; and baseline ALT or AST >3× the upper limit of normal.

### *NAT2* genotyping

Oral swab was collected at enrollment for genotyping. At the Campo Grande site, *NAT2* genotyping for acetylator-phenotype allocation was performed by long-read amplicon sequencing on the Oxford Nanopore MinION platform using a previously described multiplex pharmacogenomic panel that targets seven clinically relevant *NAT2* single-nucleotide polymorphisms (SNPs) [16]. At the Manaus site, participants were first screened using TaqMan SNP genotyping assays (Thermo Fisher Scientific) covering four canonical *NAT2* variants (four canonical NAT2 variants; Supplementary Methods); results were confirmed by Nanopore sequencing. Acetylator phenotype (slow, intermediate, or rapid) was assigned from phased *NAT2* haplotypes using the international consensus nomenclature for arylamine N-acetyltransferases [17].

### Allocation and dosing

Participants were allocated to dosing groups based on NAT2 phenotype and received once-weekly 3HP; rifapentine was dosed at 15 mg/kg (maximum 900 mg) throughout with pyridoxine 50 mg weekly. On Day 0, all participants received standard INH 15 mg/kg (maximum 900 mg). On Day 7, intermediate acetylators continued at the standard dose, rapid acetylators received an increased dose of 25 mg/kg (maximum 1,500 mg), and slow acetylators received a reduced dose of 5 mg/kg (maximum 300 mg). This conservative dose was chosen to prioritize safety before characterizing INH kinetics at the higher weekly dose. Because weight-based targets exceeded dose caps for most participants, the genotype-guided day functioned as fixed dosing: 1, 3, and 5 tablets of 300 mg INH for slow, intermediate, and rapid acetylators. From Day 14 onward, all participants received the standard 15 mg/kg dose for the remaining 10 weeks. The first three doses were directly observed; subsequent doses were self-administered with weekly telephone follow-up. Participants deprived of liberty received all doses under direct observation.

### Pharmacokinetic sampling and drug quantification

Serial PK sampling was performed after the Day 7 (genotype-guided) dose and the Day 14 (standard reference) dose. Venous blood samples were collected at 1, 2, 8, and 24 hours post-dose processed within 30 minutes, and stored at −80°C. Plasma concentrations of INH and N-acetyl-isoniazid were quantified by a validated LC-MS/MS assay at the Center for Discovery & Innovation, New Jersey, USA. LC-MS/MS assay details and performance characteristics are provided in the Supplementary Methods; the lower limit of quantification was 0.01 µg/mL.

### Population pharmacokinetic modeling

Population PK analyzes were conducted using non-linear mixed-effects modeling in the nlmixr2 (version 5.0.2) and rxode2 R packages, with parameters estimated by first-order conditional estimation with interaction (FOCEI)[18]. We evaluated one-and two-compartment disposition structures with first-order absorption and either linear or saturable (Michaelis-Menten) elimination, and additive, proportional, and combined residual-error models.

Observations below the lower limit of quantification (39 of 816; 4.8%) were retained and handled by Beal’s M3 censored-likelihood method. Allometric scaling of disposition parameters by body weight was applied a priori using fixed exponents of 0.75 on the maximum elimination rate (V_max_) and inter-compartmental clearance (Q) and 1.0 on the central (V_2_) and peripheral (V_3_) volumes (reference weight 70 kg). The effect of NAT2 acetylator phenotype was incorporated as a structural covariate on V_max_ (additive on the log scale), with intermediate acetylators as the reference group; the Michaelis constant K_m_ was shared across phenotypes. Covariates were assessed by forward inclusion (P < 0.05) and backward elimination (P < 0.01). Model selection was based on the objective function value, parameter precision, goodness-of-fit diagnostics, and visual predictive checks. The final model was a two-compartment model with first-order absorption and saturable NAT2-mediated (Michaelis-Menten) elimination from the central compartment, with allometric scaling and acetylator-stratified V_max_. Confidence intervals are reported in Table S2. Per-subject AUC0–24, C□□□, and C24 were estimated by noncompartmental analysis; a model-based sensitivity analysis is reported in Table S5. Additional methodological details, including model selection criteria, goodness-of-fit diagnostics, and confidence interval derivation, are provided in the Supplementary Methods.

### Dose-finding simulation

Phenotype-specific isoniazid dose recommendations were derived by Monte Carlo simulation from the final Michaelis-Menten popPK model. For each phenotype, 2,000 virtual subjects (weights resampled from observed cohort) were simulated across 200–1,800 mg in 50 mg steps. AUC0–24 and 24-hour post-dose concentration (C24) were derived from dense concentration profiles by trapezoidal integration. The reference target was the observed median AUC0–24 in intermediate acetylators at 900 mg flat dose (39.2 mg·h/L; IQR 23.7–50.7). The AUC-matching dose minimized |simulated AUC0–24 – reference|; target attainment and C24 exceedance were also reported. Additional simulation details, including virtual subject parameterization and target attainment criteria, are provided in the Supplementary Methods.

### Outcomes and statistical analysis

The primary outcome was pharmacokinetic similarity (no statistically significant difference in AUC0–24) between genotype-guided dosing in slow or rapid acetylators and standard 15 mg/kg dosing in intermediate acetylators. Secondary PK parameters were C□□□ and C24. The primary comparison between genotype-guided and standard dosing in intermediate acetylators was performed using the two-sided Wilcoxon rank-sum test; a non-significant result was interpreted as indicating no statistically significant difference in AUC0–24 between the groups (pharmacokinetic similarity). The study was powered at 90% to detect a one-standard-deviation difference in AUC0–24 between groups. Three pre-specified secondary comparisons were performed: (i) within-subject comparison of AUC0–24 under genotype-guided versus standard dosing in slow and rapid acetylators using the Wilcoxon signed-rank test for paired data; (ii) between-group comparison of AUC0–24 under standard dosing across acetylator phenotypes using the Wilcoxon rank-sum test; and (iii) comparison of the proportion of participants with C24 exceeding the 0.15 µg/mL threshold previously associated with systemic drug reactions during 3HP[11] using McNemar’s chi-square test with continuity correction.

## Results

### Study population and NAT2 acetylator status

Between March 2023 and December 2025, 701 individuals were assessed for eligibility across the two sites (492 at Campo Grande; 209 at Manaus), of whom 228 met inclusion criteria and were enrolled: 163 (71.5%) at Campo Grande and 65 (28.5%) at Manaus (Table S1). The enrolled cohort was 66.2% male (151/228), with a median age of 39 years (IQR 31-47) and a median weight of 72.0 kg (IQR 63.1-84.0). NAT2 nanopore sequencing classified 108 participants (47.4%) as intermediate acetylators, 99 (43.4%) as slow acetylators, and 21 (9.2%) as rapid acetylators. Four of 61 Manaus participants with both TaqMan and nanopore results were discordant (slow by TaqMan, intermediate by nanopore); these received the slow-acetylator dose. One underwent PK sampling but was excluded from the PK analysis due to dose-phenotype mismatch.

Of the 228 genotyped participants, 104 (45.6%) completed PK sampling at 1, 2, 8, and 24 hours post-dose following both the Day 7 (genotype-guided) and Day 14 (standard 900 mg flat) doses and constituted the popPK analysis set (Table 1). The subset comprised 45 intermediate, 43 slow, and 16 rapid acetylators (one slow acetylator excluded from paired analyzes due to incomplete sampling).

**Table 1.**
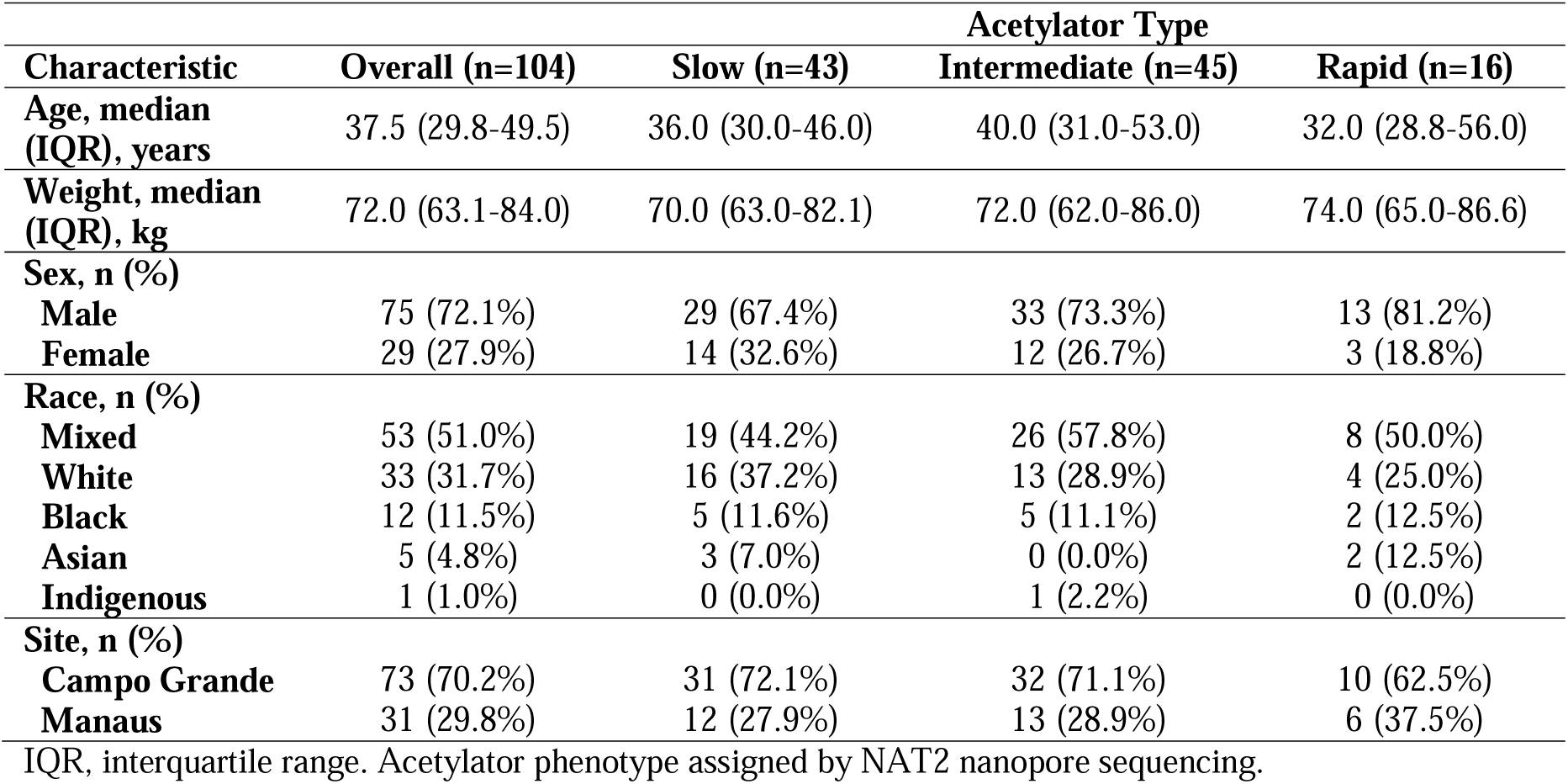
Baseline characteristics of participants with intensive PK sampling (n = 104) . Subset of participants who completed intensive sampling at 1, 2, 8 and 24 h post-dose on Day 7 (genotype-guided) and Day 14 (flat 900 mg).

### Population pharmacokinetics of isoniazid and the effect of NAT2 on clearance

The final popPK model was fit to 816 plasma isoniazid concentrations from the intensively sampled participants (Table S2). A two-compartment model with first-order absorption and saturable Michaelis-Menten elimination from the central compartment fit the databetter than thelinear-clearance alternative (ΔAIC = −121; Table S3). The Michaelis constant K_m_ was 7.45 µg/mL (RSE 27%) and was shared across phenotypes. Visual predictive checks showed concordance between observed and model-predicted median and 5th–95th-percentile profiles within each phenotype (Figure S1), and goodness-of-fit diagnostics showed predictions largely unbiased, with conditional weighted residuals centered on zero across the sampling window (Figure S2).

### Genotype-guided dosing harmonized isoniazid exposure across NAT2 phenotypes

Plasma INH concentration-time profiles differed markedly between the genotype-guided and the standard 900 mg flat-dose occasions, consistent with the underlying acetylator-stratified clearance (Figure 1; individual profiles in Figure S3). Under the standard 900 mg dose, slow acetylators showed sustained elevated INH concentrations across the dosing interval, while rapid acetylators showed lower peak and 24-hour post-dose concentrations, consistent with subtherapeutic exposure. On the genotype-guided day, slow acetylators receiving 5 mg/kg (300 mg) had a 3.7-fold reduction in median AUC0–24 (18.9 vs 70.7 mg·h/L; paired Wilcoxon p < 0.0001), rapid acetylators receiving 25 mg/kg (1,500 mg) had a 2.1-fold increase (42.4 vs 20.2 mg·h/L; p < 0.0001), and intermediate acetylators (who received 900 mg on both occasions) showed no significant change in AUC0–24 or C□□□ (33.0 vs 39.2 mg·h/L; p = 0.25), (Table S4). Across the full PK cohort, the AUC0–24 distribution was narrower under genotype-guided dosing (2.2-fold spread across phenotype medians: 18.9/33.0/42.4 mg·h/L) than under the standard 900 mg dose (3.5-fold: 20.2/39.2/70.7 mg·h/L; Figure 2). In the primary between-group comparison, guided-dose rapid acetylators showed no significant difference in AUC0–24 compared with standard-dose intermediate acetylators [42.4 (29.0–63.8) vs. 39.2 (23.7–50.7) mg·h/L; Wilcoxon rank-sum p = 0.38; median ratio 1.08], supporting pharmacokinetic similarity of the 1,500 mg guided dose. Guided-dose slow acetylators, however, fell below the reference [18.9 (11.9–24.8) vs. 39.2 (23.7–50.7) mg·h/L; p < 0.001; median ratio 0.48], confirming that the 5 mg/kg (300 mg) dose under-shot the exposure target in slow acetylators.

**Figure 1.**
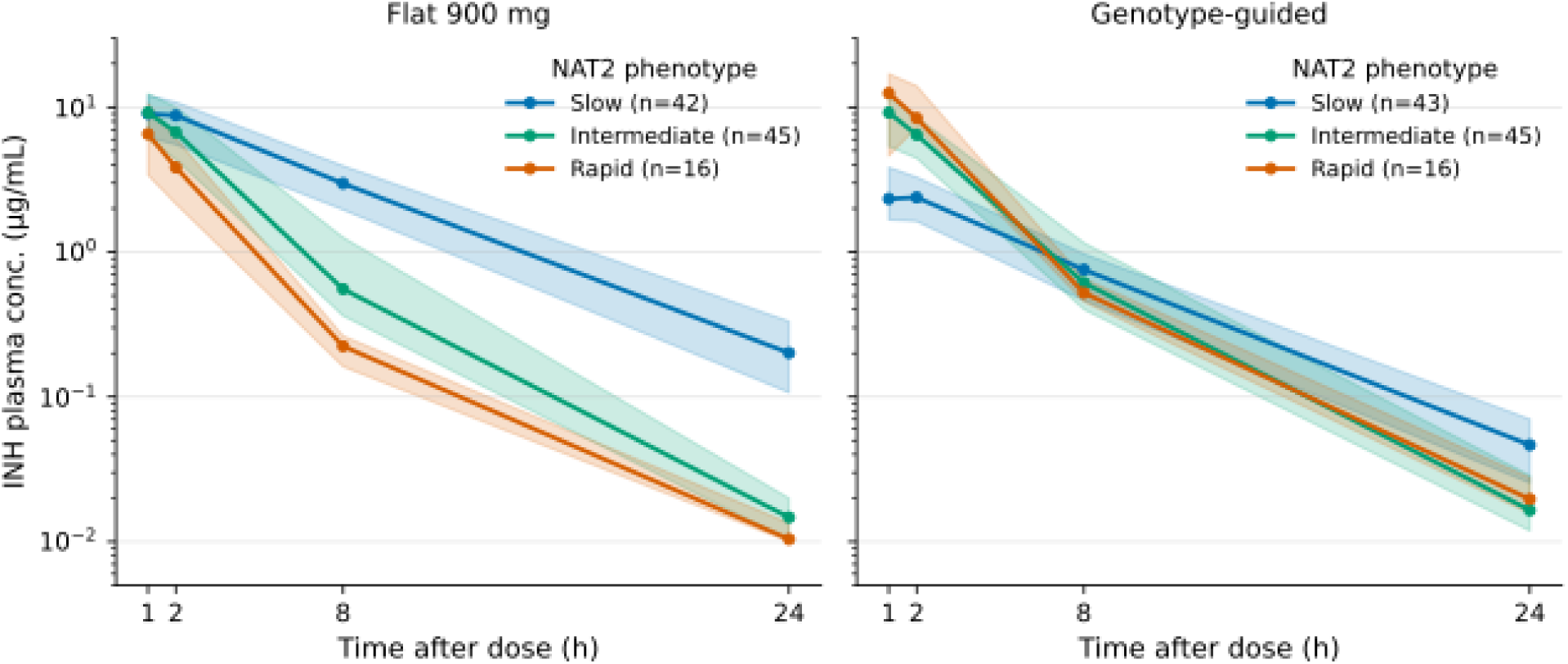
Isoniazid concentration-time profiles by dosing strategy and NAT2 acetylator phenotype. Left panel: flat 900 mg dose. Right panel: genotype-guided dose (slow, 5 mg/kg; intermediate, 15 mg/kg; rapid, 25 mg/kg; capped at 300 mg in slow, 900 mg in intermediate, and 1,500 mg in rapid acetylators). Solid lines connect median concentrations at each nominal sampling time (1, 2, 8 and 24 h post-dose); shaded ribbons denote the interquartile range.

**Figure 2.**
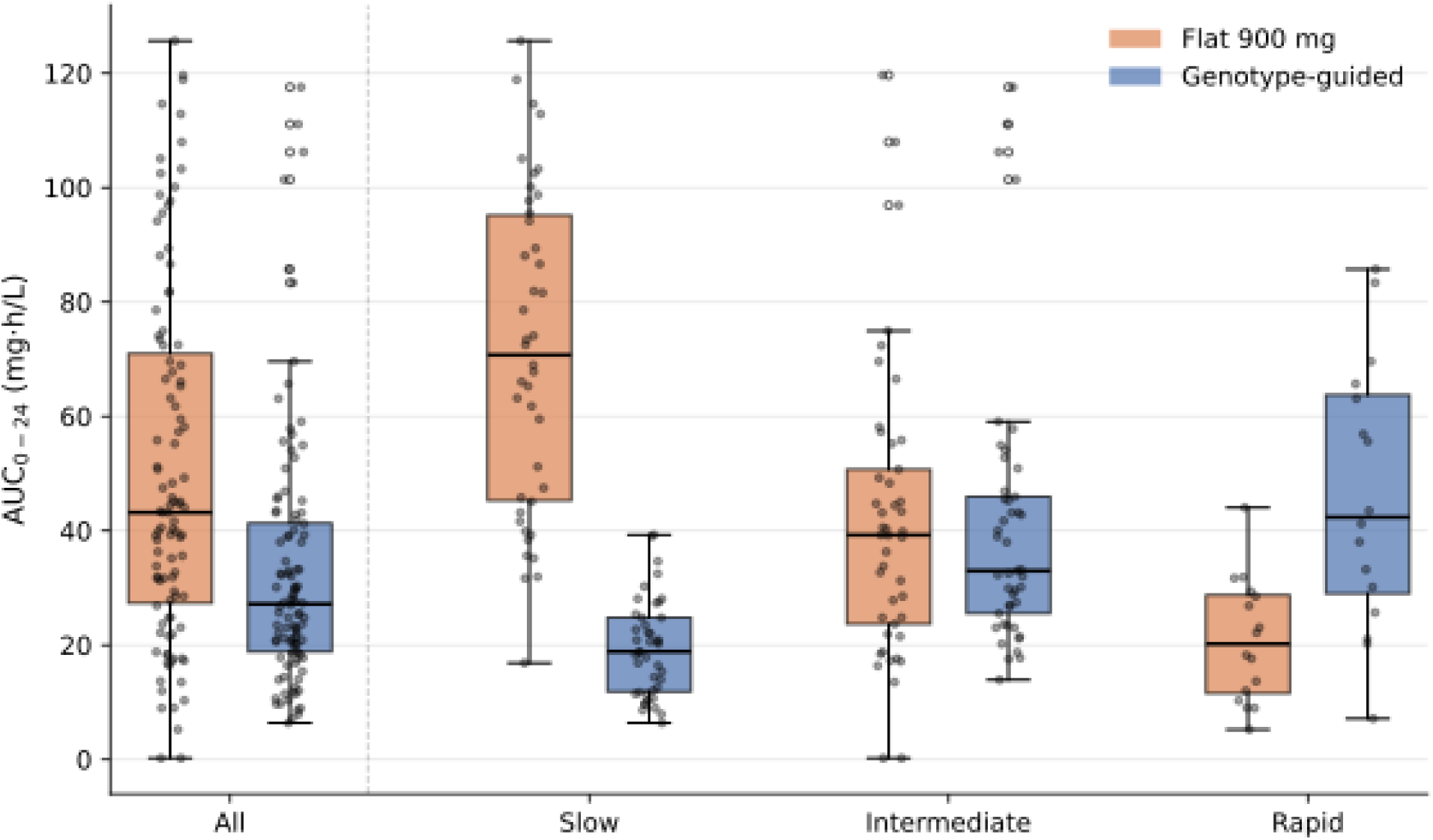
Isoniazid AUC0–24 by NAT2 acetylator phenotype and dosing strategy. Box-and-whisker plots of the area under the plasma isoniazid concentration–time curve from 0 to 24 hours (AUC0–24) in the 103 participants who completed PK sampling on both occasions. The leftmost pair of boxes (All, n=103) shows the overall distribution across all phenotypes combined; subsequent pairs are stratified by NAT2 acetylator phenotype (slow n=42, intermediate n=45, rapid n=16). Within each group, the left (orange) box corresponds to the flat 900 mg occasion (Day 14) and the right (blue) box to the genotype-guided occasion (Day 7). Boxes denote the interquartile range, the horizontal line within each box the median, whiskers the 1.5 × IQR rule, and individual points the observed per-subject values. Paired within-subject comparisons and Wilcoxon p-values are reported in Table S4.

Maximum concentration (C□□□) followed the same pattern; under genotype-guided dosing, slow acetylators had a 3.6-fold lower C□□□ (2.6 vs 9.3 µg/mL; p < 0.0001), rapid acetylators a 2.0-fold higher C□□□ (13.3 vs 6.5 µg/mL; p < 0.001), and intermediate acetylators no significant change (9.5 vs 9.8 µg/mL; p = 0.29) (Table S4). Within-subject changes between the standard 900 mg and genotype-guided occasions confirmed this harmonization (Figure 3). AUC□□□□ decreased in 41/42 slow acetylators (median reduction 74%) and increased in 15/16 rapid acetylators (median increase 126%) on the genotype-guided occasion. Intermediate acetylators showed no systematic change, as expected. The observed Cmax IQR across all participants under the standard 900 mg dose was 6.2–12 µg/mL, spanning the estimated Michaelis constant (Km = 7.45 µg/mL), consistent with partial NAT2 saturation.

**Figure 3.**
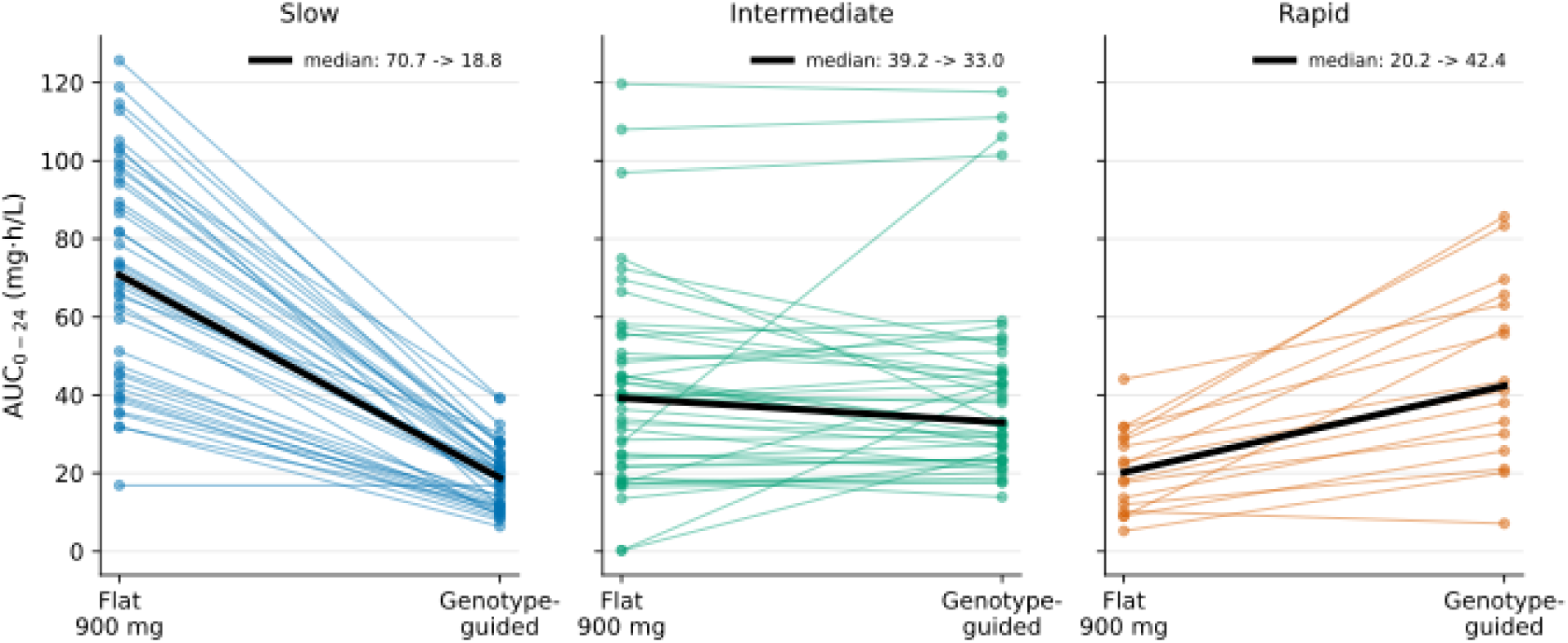
Within-subject change in isoniazid AUC0–24 between the flat 900 mg and genotype-guided doses, stratified by NAT2 acetylator phenotype. Paired AUC0–24 values for each of the participants are connected by a thin colored line between the flat 900 mg occasion (Day 14, left position in each panel) and the genotype-guided occasion (Day 7, right position). Panels are stratified by NAT2 acetylator phenotype (slow, n = 42; intermediate, n = 45; rapid, n = 16). The thick black line overlay shows the within-phenotype median trajectory, with the corresponding median values annotated. Genotype-guided dosing produces a large reduction in median AUC0–24 in slow acetylators (70.7 → 18.9 mg·h/L), a small non-significant change in intermediate acetylators (39.2 → 33.0 mg·h/L; Wilcoxon signed-rank p = 0.25), and an increase in rapid acetylators (20.2 → 42.4 mg·h/L).

### Genotype-guided dosing reduced supratherapeutic 24-hour post-dose concentrations

Under standard 900 mg flat dosing, slow acetylators had a median 24-hour post-dose concentration (C24) roughly 20-fold higher than that of intermediate acetylators (0.20 vs 0.01 µg/mL; Figure 4), reflecting sustained supratherapeutic INH exposure between weekly doses. With genotype-guided dose reduction to 5 mg/kg, median C24 in slow acetylators fell to 0.05 µg/mL (paired Wilcoxon p < 0.0001; Table S4). Among the slow acetylators with paired sampling on both occasions, the proportion with C24 above the 0.15 µg/mL threshold previously associated with systemic drug reactions during 3HP[11] fell from 27/42 (64%) on the flat 900 mg dose to 1/42 (2%) on the genotype-guided dose (McNemar χ² = 24.0, p < 0.0001; Table S4 and Figure 4). Results were robust to exposure-estimation method (model-based vs. non-compartmental; Table S5).

**Figure 4.**
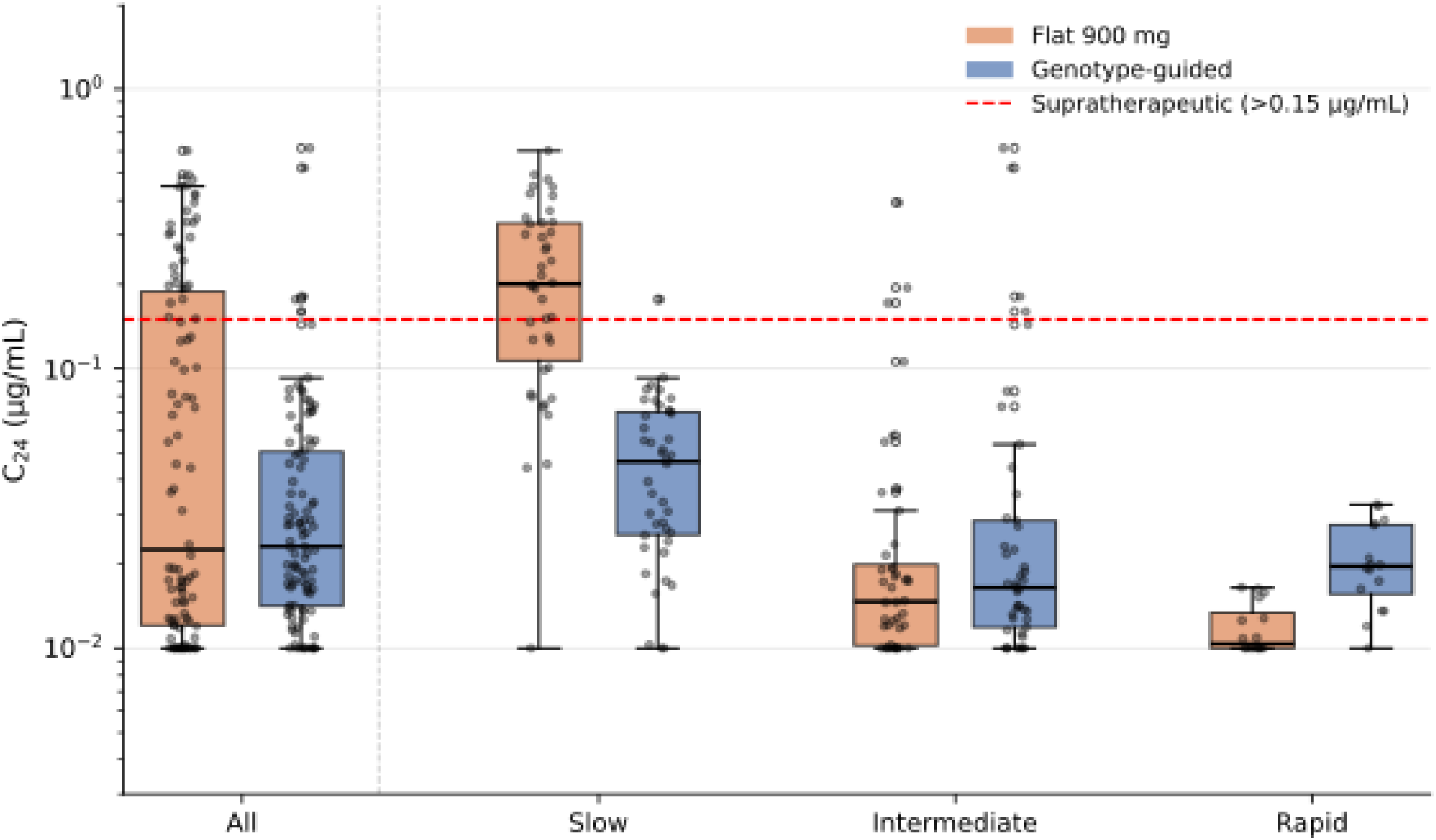
Isoniazid 24-hour post-dose plasma concentration (C_24_) by NAT2 acetylator phenotype and dosing strategy. Box-and-whisker plots of the 24-hour post-dose plasma isoniazid concentration (C24) in the 102 participants with a 24-hour sample on both occasions. The leftmost pair (All, n=102) shows the overall distribution across all phenotypes; subsequent pairs are stratified by NAT2 acetylator phenotype (slow n=42, intermediate n=44, rapid n=16). Within each group, the left (orange) box corresponds to the flat 900 mg occasion and the right (blue) box to the genotype-guided occasion. The y-axis is on a logarithmic scale to accommodate the wide between-subject dispersion in concentrations. The dashed red horizontal line marks the prespecified supratherapeutic threshold of 0.15 µg/mL. Box, median, whisker, and point conventions are identical to those of Figure 2. Boxes denote the interquartile range, the horizontal line within each box the median, whiskers the 1.5 × IQR rule, and individual points the observed per-subject values. Values at 0.01 µg/mL correspond to the lower limit of quantification (BLQ = 0.01 µg/mL); concentrations below this threshold were not quantifiable.

### Acetylator type-specific isoniazid dose recommendations

Monte Carlo simulations identified phenotype-specific doses predicted to match the intermediate-acetylator AUC0–24 at the standard 900 mg dose. AUC-matching doses (AUC0–24 closest to the 39.2 mg·h/L reference) were 600 mg in slow acetylators, 950 mg in intermediate acetylators, and 1,300 mg in rapid acetylators. Rounding to 300 mg tablet increments yielded final recommendations of 600/900/1,200 mg for slow/intermediate/rapid acetylators (Figure 5). At the recommended doses, the median simulated AUC0–24 was 38.7 mg·h/L in slow acetylators (76.3% of subjects within the reference interquartile range), 36.0 mg·h/L in intermediate acetylators (74.2%), and 35.7 mg·h/L in rapid acetylators (69.6%). The proportion of subjects with C24 exceeding 0.15 µg/mL was 0.8% (intermediate) and 0.0% (rapid). In slow acetylators, the 600 mg dose was associated with a higher C24 exceedance (31.6%) than in the other phenotypes, reflecting the structurally lower elimination capacity in this group. No dose simultaneously achieved the AUC-matching target and the 15% C24 supratherapeutic threshold for slow acetylators (the dose required to bring C24 exceedance below 15% was 400 mg, delivering a median AUC0–24 of only 22.9 mg·h/L, approximately 59% of the reference target).

**Figure 5.**
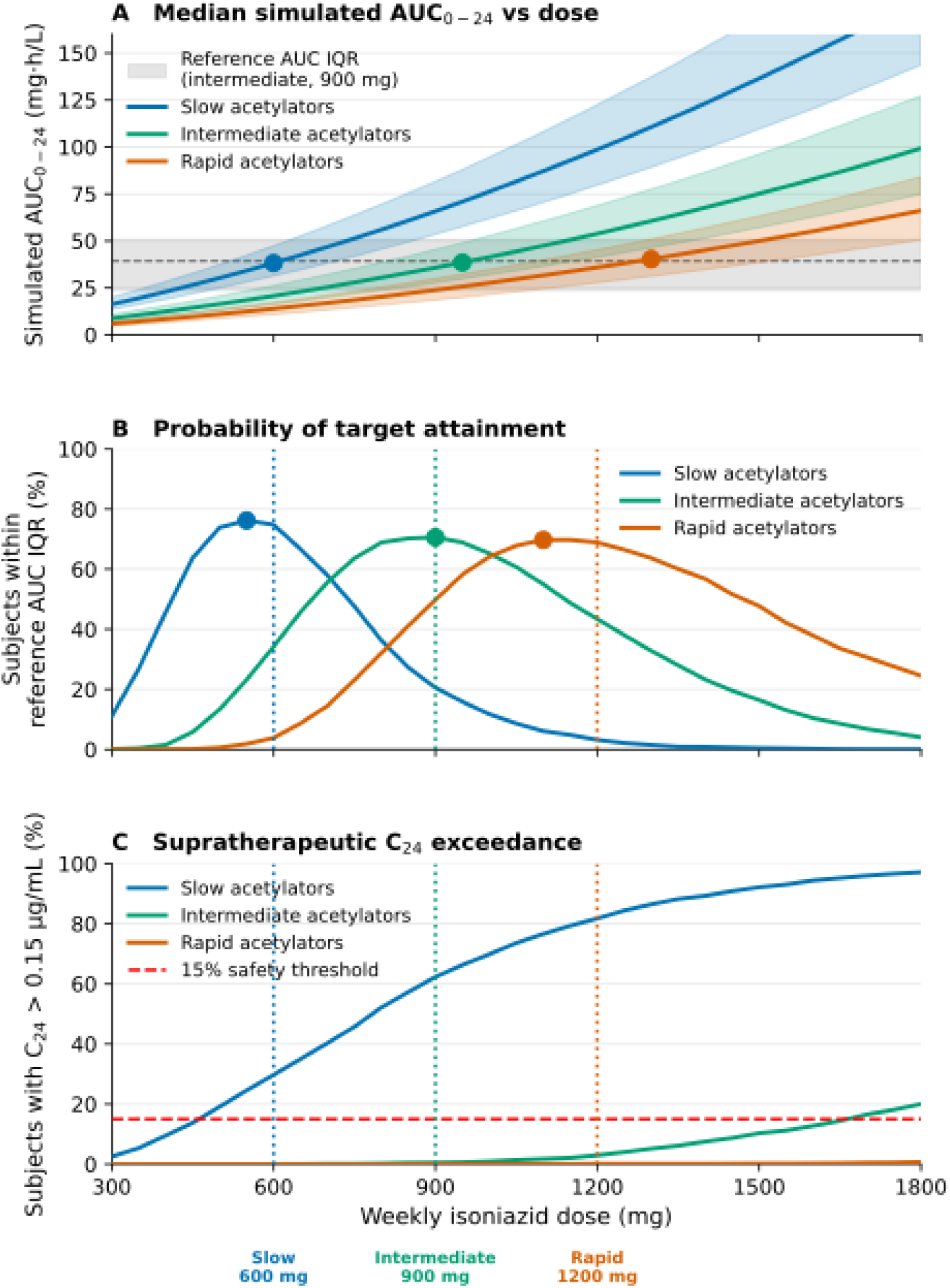
Phenotype-specific isoniazid dose recommendations from Monte Carlo simulation of the final Michaelis-Menten popPK model. (A) Median simulated AUC□□□□ (line) with interquartile range (shaded band) versus weekly dose; the grey horizontal band is the intermediate-acetylator reference exposure IQR at the standard 900 mg dose, and filled circles mark each phenotype’s AUC-matching dose (the dose whose median simulated AUC□□□□ is closest to the 39.2 mg·h/L intermediate-acetylator reference; a horizontal reference line at 39.2 mg·h/L is shown for comparison). (B) Probability of target attainment, defined as the percentage of simulated subjects whose AUC□□□□ falls within the intermediate-900 mg reference IQR, versus weekly dose; filled circles mark each phenotype’s dose maximizing the percentage within the reference IQR, and dotted vertical lines indicate the final rounded recommended doses (600, 900, 1,200 mg for slow, intermediate, rapid acetylators, respectively). (C) Proportion of simulated subjects with C24 above the 0.15 µg/mL supratherapeutic threshold versus weekly dose; the red dashed line marks the 15 % safety threshold used in dose selection.

## Discussion

In this prospective pharmacogenomic-guided dosing study, we demonstrated that NAT2 genotype remains a major determinant of INH pharmacokinetics during TPT with 3HP and that genotype-guided dose adjustment substantially reduces interindividual variability in drug exposure. Slow acetylators had approximately half the clearance of intermediate acetylators; rapid acetylators had the highest, resulting in marked exposure differences under standard dosing. By reducing the INH dose in slow acetylators and increasing it in rapid acetylators, we narrowed exposure across acetylator phenotypes and reduced prolonged supratherapeutic concentrations. These findings provide guidance for NAT2-guided dosing for the weekly high-dose INH regimen used in 3HP.

Our findings extend prior work establishing NAT2 as the principal determinant of isoniazid metabolism and toxicity.[10, 19, 20] Early PK studies showed substantially higher exposure in slow versus rapid acetylators at identical doses.[21, 22] The randomized trial by Azuma and colleagues demonstrated that *NAT2*-guided daily dose adjustment reduced both INH-induced liver injury and early treatment failure during four-drug TB therapy.[14] Earlier work testing NAT2-guided INH dosing in daily therapy[23] and a pilot study of NAT2-guided INH monotherapy similarly normalized exposure in slow acetylators.[15] Our results extend these findings to 3HP, confirming that NAT2-driven clearance differences persist at the higher weekly dose and can be corrected by phenotype-specific adjustment.

A key finding was the magnitude of exposure heterogeneity under standard dosing. Slow acetylators had more than three-fold higher exposure than rapid acetylators and frequently maintained elevated 24-hour post-dose concentrations. Genotype-guided dosing narrowed the exposure distribution, bringing slow and rapid acetylators closer to the intermediate range. The intervention reduced excessive exposure in slow acetylators while increasing it in rapid acetylators, who are at risk of subtherapeutic levels.[24] Reducing prolonged exposure among slow acetylators may improve 3HP safety.[25] Previous studies have identified both NAT2 slow-acetylator status and elevated INH concentrations as predictors of systemic drug reactions during weekly rifapentine-INH therapy.[11, 26, 27] In our study, the proportion of participants exceeding the previously reported C24 threshold associated with systemic drug reactions[11] decreased from 64% under standard dosing to 2% under genotype-guided dosing. Although not powered for clinical adverse events, these findings suggest that some toxicity risk may be predictable and modifiable through pharmacogenomic-guided dosing.

Our use of a saturable, two-compartment model is consistent with the enzymology of isoniazid, which is eliminated principally by capacity-limited NAT2 acetylation and therefore follows Michaelis-Menten rather than strictly first-order kinetics.[28, 29] Because the estimated Km (7.5 µg/mL) lay within the observed Cmax range, acetylation was partially saturated at weekly-dose concentrations, so linear clearance no longer holds. This explains why prior isoniazid popPK models developed at standard daily doses adequately used linear clearance with NAT2 as a covariate,[21] whereas the higher weekly doses and intensive sampling in this study revealed the underlying nonlinearity. Modeling acetylator phenotype as a shift in Vmax with a shared Km is also biologically grounded, since slow-acetylator NAT2 variants reduce enzyme capacity rather than substrate affinity.[7, 21, 22]

Simulations from the final popPK model yielded phenotype-specific dose recommendations of 600, 900, and 1,200 mg for slow, intermediate, and rapid acetylators, approximating the reference AUC0–24 while remaining compatible with 300 mg tablets. Because NAT2-mediated acetylation is partially saturated at clinical INH concentrations (estimated Km 7.5 µg/mL, within the observed Cmax IQR of 6.2–12 µg/mL), these recommendations reflect Michaelis–Menten rather than linear elimination. Linear extrapolation overpredicts the rapid-acetylator dose (1,500 mg protocol vs. 1,200 mg simulated) because Vmax is high but finite. The recommended 600 mg dose in slow acetylators represents a trade-off: no dose simultaneously achieved the AUC target and the 15% C24 threshold because of their lower elimination capacity and the short weekly dosing interval. The 0.15 µg/mL C24 threshold was derived from a single observational 3HP cohort and has not been prospectively validated as a safety target.[11] The balance between exposure adequacy and toxicity in slow acetylators should be re-examined in trials with clinical endpoints.

These dose recommendations are specific to once-weekly 3HP and should not be extrapolated to 1HP.[30, 31] Under 1HP, daily INH doses (300 mg) produce peak concentrations well below the estimated Km (7.45 µg/mL), placing acetylation in a near-linear rather than saturable regime;[22] our phenotype-specific adjustments therefore do not apply directly. NAT2 genotype is nonetheless expected to influence cumulative INH exposure and hepatotoxicity risk during 1HP,[10] warranting dedicated pharmacokinetic study. A further consideration for implementation is cost. Genotype-guided dosing requires an upfront NAT2 assay, and the long-read nanopore sequencing and TaqMan platforms used here, while accurate, remain relatively expensive and infrastructure-dependent for many high-burden, resource-limited settings. Formal cost-effectiveness analysis and development of simpler point-of-care assays will be needed to establish whether NAT2-guided dosing can be delivered affordably at scale.

Our findings should be interpreted within the context of some limitations. First, the study was designed as a PK trial and was not powered to evaluate clinical outcomes such as TB prevention efficacy, treatment completion, hepatotoxicity, or systemic drug reactions. Reduced supratherapeutic exposure should be interpreted as reduction in a validated PK risk marker, rather than direct evidence of improved safety. Second, the number of rapid acetylators was small, resulting in less precise clearance estimates in this subgroup. Third, the absorption rate constant was unidentifiable from the sparse early sampling, limiting characterization of the absorption phase. Fourth, dosing was evaluated during a single week; longer-term PK adaptation or adherence factors could affect full-course exposure. Fifth, the phenotype-specific dose recommendations are based on simulations from a single cohort and require prospective validation, ideally with clinical endpoints. Finally, additional studies are needed to assess generalizability across populations with different NAT2 allele frequencies.

In conclusion, NAT2 genotype strongly influences INH exposure during once-weekly 3HP therapy, resulting in substantial pharmacokinetic heterogeneity under standard dosing. A simple genotype-guided dosing strategy reduced excessive exposure among slow acetylators, increased exposure among rapid acetylators, and harmonized pharmacokinetic profiles across acetylator phenotypes. These findings support further evaluation of NAT2-guided precision dosing as a strategy to improve the safety and consistency of TPT and provide a framework for integrating pharmacogenomics into large-scale TB prevention programs.

## Acknowledgments

The authors thank the study participants and the clinical teams at UFMS (Campo Grande) and FMT-HVD (Manaus) for their contribution to this work.

## Author contributions

K.E.S: Conceptualization, Data curation, Formal analysis, Methodology, Visualization, Writing – original draft, Writing – review & editing. S.S.: Formal analysis, Writing – review & editing. K.M, A.G.C: Investigation, Data curation, Project administration, Writing – review & editing. P.V, R.D.O, M.A.M.P, R.S.G, J.P.G.M: Investigation, Writing – review & editing. P.C.P, E.Y: Data curation, Writing – review & editing. R.S, M.C.-S.: Conceptualization, Resources, Supervision, Writing – review & editing. J.C, J.R.A.: Conceptualization, Funding acquisition, Methodology, Project administration, Resources, Supervision, Writing – review & editing.

## Data Availability

Deidentified individual participant pharmacokinetic data, metadata, and analysis code are publicly available at https://github.com/kesiaeds/POCPOI-NAT2-isoniazid-PK. Supplementary Methods, 4 Supplementary Figures (S1–S4), and 5 Supplementary Tables (S1–S5) are provided as a single supplementary file.

