## Supplemental Data 1 for "Genotype-guided isoniazid dosing harmonizes drug exposure in 3HP tuberculosis preventive therapy"

Supplementary Methods	2

Supplementary Figure S1	5

Supplementary Figure S2	6

Supplementary Figure S3	7

Supplementary Figure S4	8

Supplementary Table S1	9

Supplementary Table S2	10

Supplementary Table S3	11

Supplementary Table S4	12

Supplementary Table S5	13

### **Supplementary Methods**

**NAT2 Genotyping**

At the Campo Grande site, NAT2 acetylator phenotype was determined by long-read amplicon sequencing on the Oxford Nanopore Technology platform, targeting four canonical NAT2 single-nucleotide variants: rs1801279 (G191A), rs1801280 (T341C), rs1799930 (G590A), and rs1799931 (G857A)(1). Haplotype inference and phenotype assignment followed the NAT2 database nomenclature (nat.mbg.duth.gr)(2). At the Manaus site, initial phenotype assignment for dosing allocation used TaqMan qPCR targeting the same four variants; nanopore sequencing confirmation was performed subsequently for all Manaus participants with available samples.

### **Pharmacokinetic sampling and drug quantification**

Plasma isoniazid (INH) and N-acetyl-isoniazid concentrations were quantified by a validated liquid chromatography-tandem mass spectrometry (LC-MS/MS) assay performed at the Center for Discovery & Innovation (Hackensack Meridian Health, NJ, USA). Analytes were extracted from plasma by protein precipitation and detected by positive electrospray ionization with selected-reaction monitoring on an AB Sciex 5500 triple-quadrupole mass spectrometer using stable-isotope-labelled internal standards (isoniazid-d₄, N-acetyl-isoniazid-d₄). The lower limit of quantification (LLOQ) was 0.01 µg/mL for both analytes. Intra- and inter-assay precision (coefficient of variation) was ≤15% across all quality-control concentration levels. Observations below the LLOQ were flagged as below the limit of quantification (BLQ) and handled in the population PK analysis using Beal’s M3 method.

**Population pharmacokinetic model: confidence interval derivation**

For all fixed-effects parameters, 95% confidence intervals were derived from the Wald (asymptotic normal) approximation using the Fisher information matrix at the FOCEI solution. Confidence intervals for the derived baseline clearance (CL₀ = Vmax/Km) were propagated from the Wald confidence intervals of the log-scale phenotype-shift parameters using the delta method, with tVmax and tKm held at their point estimates. Parameter estimates and confidence intervals are reported in Supplementary Table S2.

**Non-compartmental analysis**

Per-subject AUC₀–₂₄, Cₘₐˣ, and C₂₄ used for inferential comparisons were computed by non-compartmental analysis using linear-trapezoidal integration, assuming C = 0 at t = 0 with linear interpolation to the 1-hour sample, applied independently to observed plasma INH concentrations at 1, 2, 8, and 24 hours post-dose on each occasion. As a sensitivity analysis, individual exposures were also re-derived from dense (1-min grid) simulations of the final Michaelis-Menten population PK model using each participant’s empirical-Bayes individual parameter estimates (MM-IPRED). Concordance between NCA and model-derived estimates is reported in Supplementary Table S5.

**Monte Carlo dose-finding simulation**

For each NAT2 phenotype, 2,000 virtual subjects were generated by resampling body weights with replacement from the observed PK cohort within phenotype, and between-subject variability in Vmax was applied on the log scale using the estimated interindividual variability from the final model. Each virtual subject was simulated across a dose grid from 200 to 1,800 mg in 50 mg increments using the rxode2 R package (3), producing dense (1-min) concentration-time profiles from which AUC₀–₂₄ and C₂₄ were derived by trapezoidal integration. The reference exposure target was the observed median AUC₀–₂₄ in intermediate acetylators at the standard 900 mg flat dose (39.24 mg·h/L; IQR 23.7–50.7 mg·h/L). The AUC-matching dose for each phenotype was defined as the dose minimizing the absolute difference between the simulated median AUC₀–₂₄ and this reference. Target attainment (proportion of virtual subjects with AUC₀–₂₄ within the reference IQR) and the proportion with C₂₄ exceeding the 0.15 µg/mL supratherapeutic threshold were also reported at each dose. Simulations were implemented in R (version 4.3) with set.seed(20260601) for reproducibility.

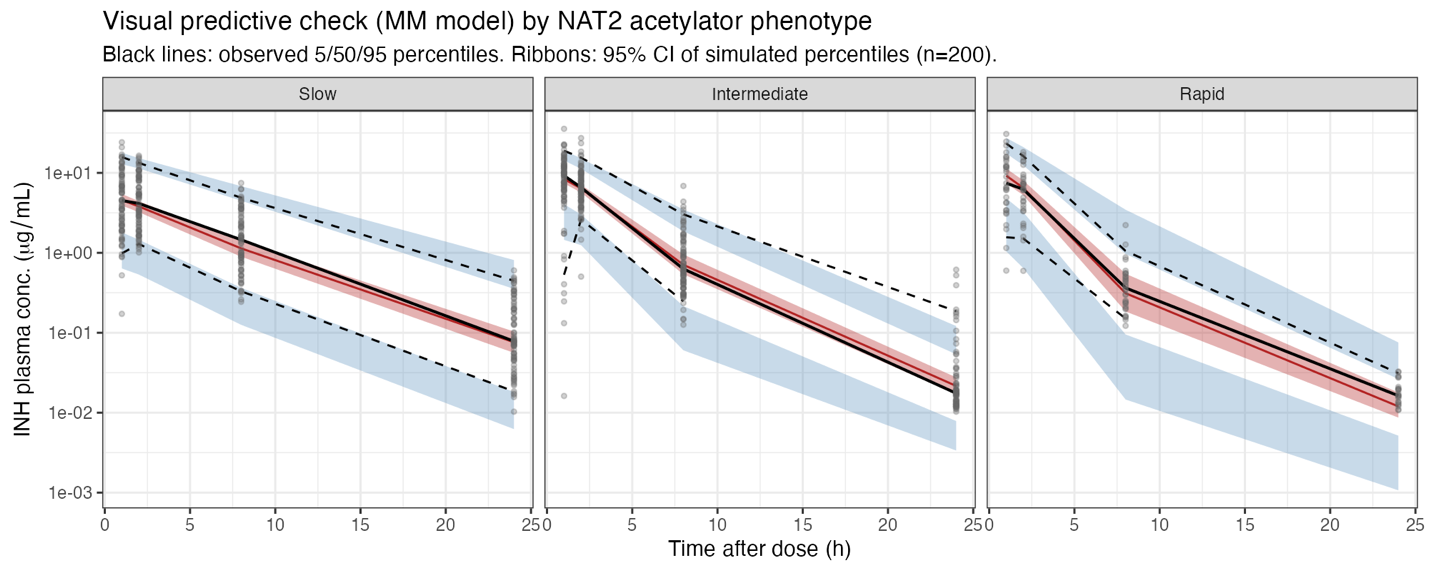

**Figure S1. Visual predictive check of the Michaelis-Menten population pharmacokinetic model, stratified by NAT2 acetylator phenotype.** Visual predictive check (VPC) based on 200 Monte Carlo replicates simulated from the final two-compartment FOCEI fit with first-order absorption and saturable Michaelis-Menten elimination from the central compartment (rxode2 ODE simulation; n = 104 subjects, 816 observations). Concentrations are stratified by nominal time-after-dose bin (1, 2, 8, 24 h). Solid black line is the observed median isoniazid concentration; dashed black lines are the observed 5th and 95th percentiles. Red ribbon and red line indicate the 95% confidence interval and median of simulated medians, respectively. Blue ribbons represent 95% confidence intervals for the simulated 5th and 95th percentiles. Grey points are individual observations.

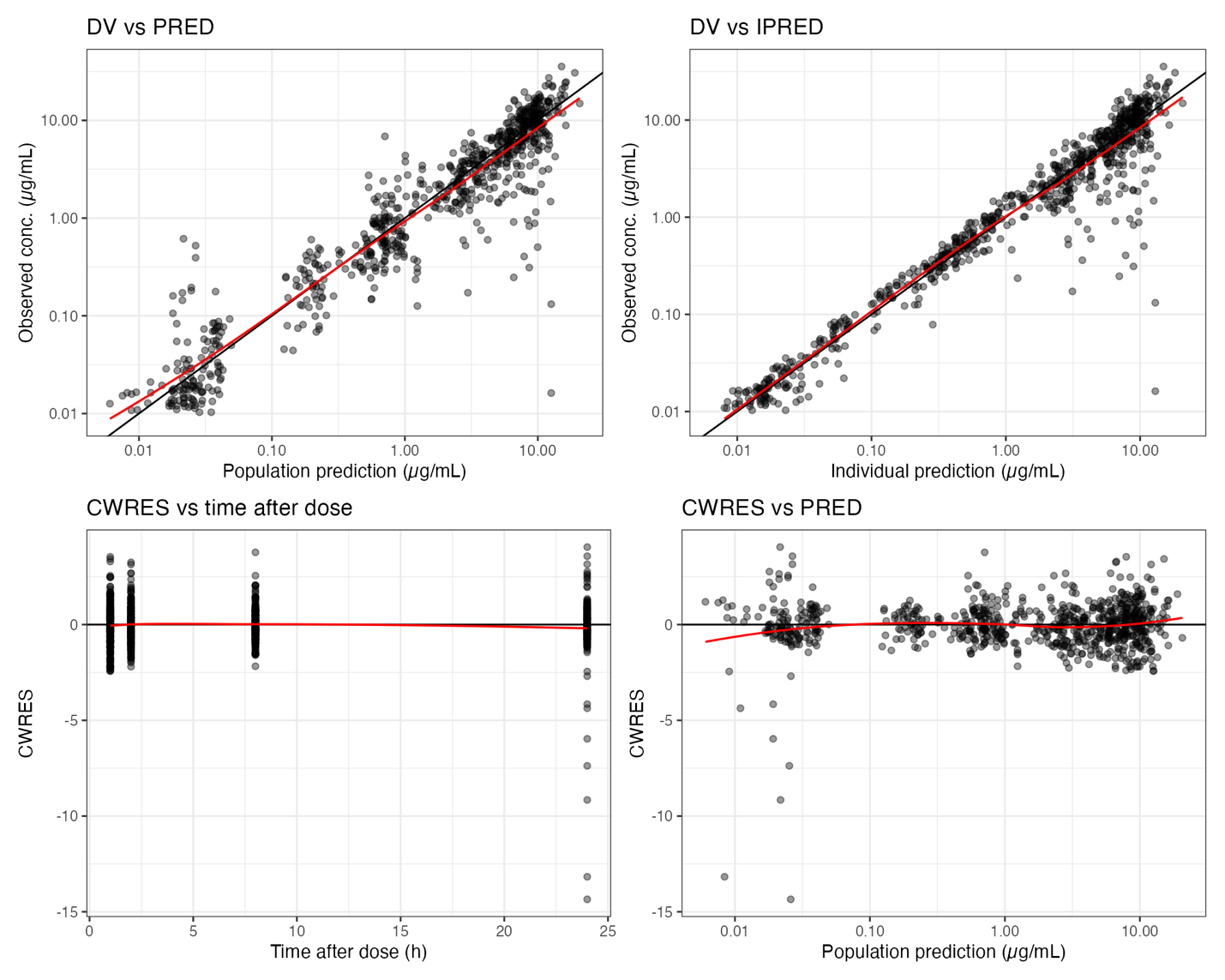

**Figure S2. Goodness-of-fit diagnostics for the final Michaelis-Menten population pharmacokinetic model.** Standard goodness-of-fit panels for the final two-compartment FOCEI fit with first-order absorption, saturable Michaelis-Menten elimination from the central compartment, and a NAT2-acetylator covariate on V_max_ (n = 104 subjects, 816 observations). Top-left: observed plasma isoniazid concentration (DV) versus population prediction (PRED) on log-log axes; black line is the line of identity, red curve is a LOESS smoother. Top-right: DV versus individual prediction (IPRED) on log-log axes with the same overlays. Bottom-left: conditional weighted residuals (CWRES) versus time after dose; black line at zero, red curve is a LOESS smoother. Bottom-right: CWRES versus PRED on log-PRED axis with the same overlays.

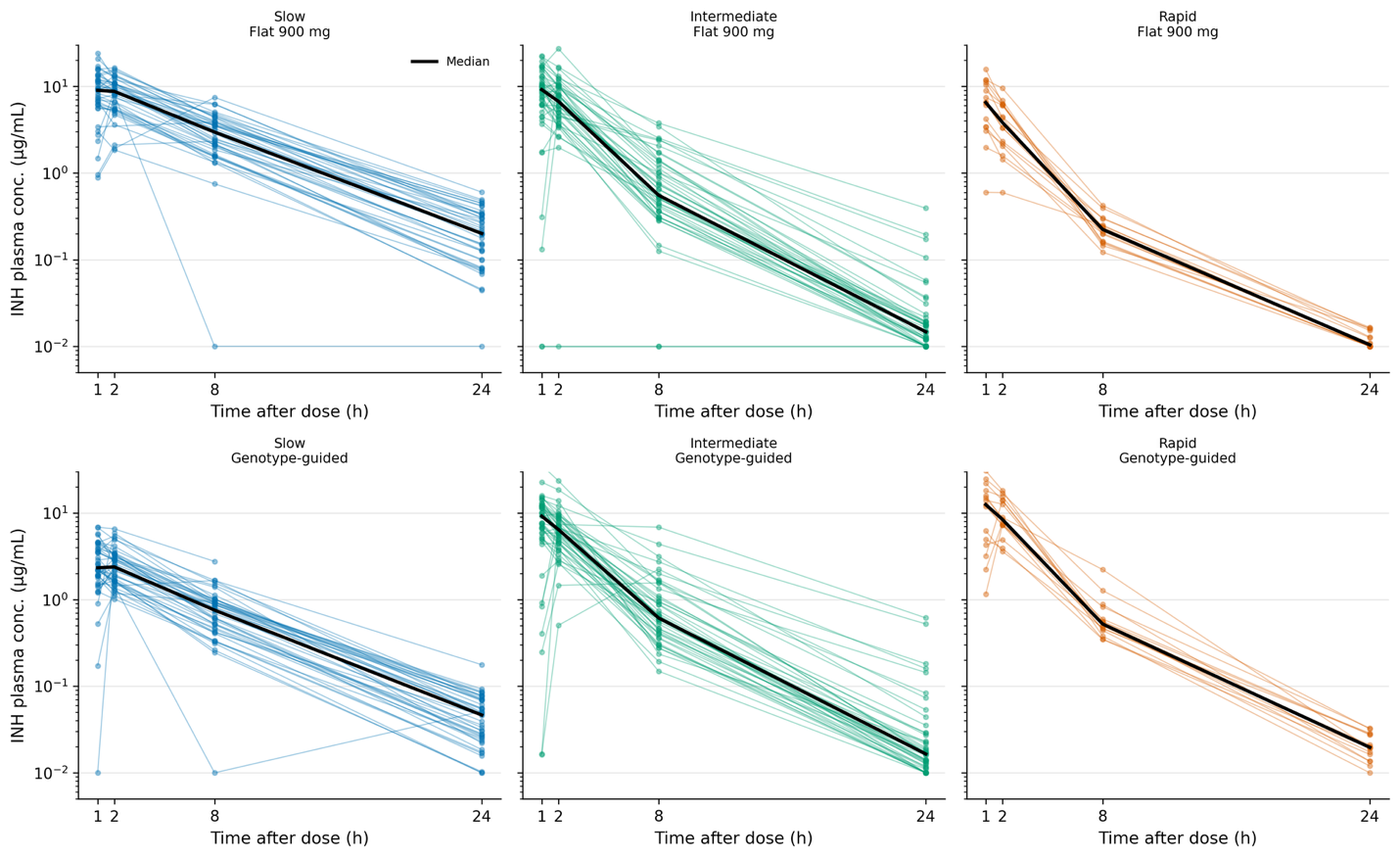

**Figure S3. Individual observed isoniazid plasma concentration-time profiles by NAT2 phenotype and dosing regimen.** Individual observed INH plasma concentrations (connected points, log scale) are shown for all participants with a pharmacokinetic profile, stratified by NAT2 acetylator phenotype (columns: slow, intermediate, rapid) and dosing occasion (rows: flat 900 mg [Day 14]; genotype-guided [Day 7]). The bold black line denotes the median profile. BLQ values (LLOQ = 0.01 µg/mL) are plotted at 0.01 µg/mL. INH, isoniazid; LLOQ, lower limit of quantification; NAT2, N-acetyltransferase 2.

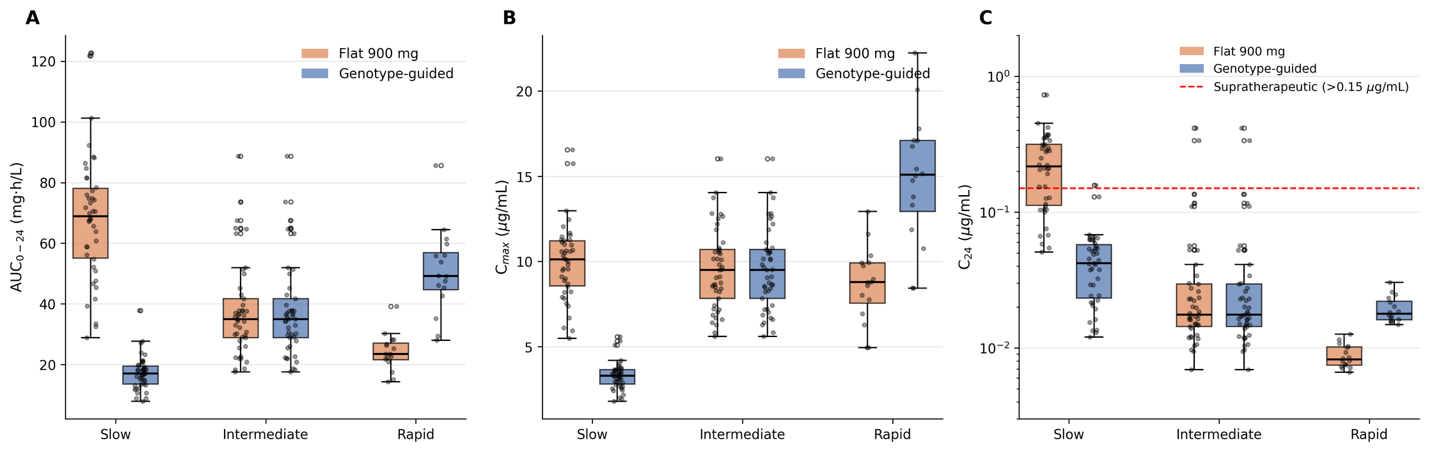

**Figure S4. Model-derived isoniazid exposures by NAT2 acetylator phenotype and dosing strategy.** Box-and-whisker plots of per-subject isoniazid exposures re-derived from the final Michaelis-Menten population pharmacokinetic model using each participant's empirical-Bayes individual parameter estimates (V_max_, K_m_, V2, Q, V3, K_a_). **(A)** Area under the plasma isoniazid concentration-time curve from 0 to 24 hours (AUC₀₋₂₄, mg·h/L). **(B)** Maximum plasma concentration (Cₘₐₓ, µg/mL). **(C)** 24-hour post-dose plasma concentration (C₂₄, µg/mL), shown on a logarithmic scale; the red dashed line marks the 0.15 µg/mL supratherapeutic threshold previously associated with systemic drug reactions during 3HP. In each panel, boxes show the median and interquartile range and whiskers extend to 1.5 × IQR; individual subject-occasion values are overlaid as jittered black points.

### **Table S1. Baseline characteristics of enrolled participants (n = 228)**

| **Characteristic** | **Overall (n=228)** | **Slow (n=99)** | **Intermediate (n=108)** | **Rapid (n=21)** |
| --- | --- | --- | --- | --- |
| **Age, median (IQR), years** | 39.0 (31.0-47.0) | 38.0 (30.0-46.0) | 40.0 (32.8-48.0) | 41.0 (29.0-50.0) |
| **Weight, median (IQR), kg** | 72.0 (63.1-84.0) | 70.0 (63.0-82.1) | 73.0 (62.3-86.0) | 74.0 (65.0-86.6) |
| **Sex, n (%)** |  |  |  |  |
| **Male** | 151 (66.2%) | 57 (57.6%) | 78 (72.2%) | 16 (76.2%) |
| **Female** | 77 (33.8%) | 42 (42.4%) | 30 (27.8%) | 5 (23.8%) |
| **Race, n (%)** |  |  |  |  |
| **Mixed** | 111 (48.7%) | 47 (47.5%) | 54 (50.0%) | 10 (47.6%) |
| **White** | 61 (26.8%) | 32 (32.3%) | 22 (20.4%) | 7 (33.3%) |
| **Black** | 45 (19.7%) | 17 (17.2%) | 26 (24.1%) | 2 (9.5%) |
| **Asian** | 10 (4.4%) | 3 (3.0%) | 5 (4.6%) | 2 (9.5%) |
| **Indigenous** | 1 (0.4%) | 0 (0.0%) | 1 (0.9%) | 0 (0.0%) |
| **Site, n (%)** |  |  |  |  |
| **Campo Grande** | 163 (71.5%) | 63 (63.6%) | 85 (78.7%) | 15 (71.4%) |
| **Manaus** | 65 (28.5%) | 36 (36.4%) | 23 (21.3%) | 6 (28.6%) |

IQR, interquartile range. Acetylator phenotype assigned by NAT2 nanopore sequencing.

**Table S2. Population pharmacokinetic parameter estimates (two-compartment model with saturable Michaelis-Menten elimination and NAT2 covariate on Vmax).** FOCEI fit (nlmixr2est 5.0.2) to 816 isoniazid plasma concentrations from the 104 intensively sampled participants. Allometric scaling fixed a priori (exponent 0.75 on V_max_ and Q, 1.0 on V_2_ and V_3_; reference weight 70 kg).

| **Parameter** | **Estimate** | **%RSE** | **95% CI** |
| --- | --- | --- | --- |
| **Maximum elimination rate, V_max_ (mg/h)** |  |  |  |
| Slow NAT2 acetylators, V_max_, slow | 159 |  | 128-197 |
| Intermediate NAT2 acetylators (reference), V_max_,inter | 300 | 17.8 | 195-405 |
| Rapid NAT2 acetylators, V_max_,rapid | 425 |  | 240-753 |
| Michaelis constant, K_m_ (µg/mL) | 7.45 | 27.3 | 3.47-11.43 |
| Apparent absorption rate, K_a_ (h⁻¹) | 6.68 | 322.8 | −35.6-48.9 |
| Apparent volume of distribution, central V_2_/F (L) | 81.8 | 6.5 | 71.4-92.2 |
| Apparent intercompartmental clearance, Q/F (L/h) | 2.22 | 50.1 | 0.04-4.40 |
| Apparent volume of distribution, peripheral V_3_/F (L) | 16.3 | 27.2 | 7.6-25.0 |
| Allometric exponent on V_max_ and Q (fixed) | 0.75 | fixed |  |
| Allometric exponent on V_2_ and V_3_ (fixed) | 1.0 | fixed |  |
| **Residual variability** |  |  |  |
| Proportional error | 39.9% |  |  |
| Interindividual variability on V_max_ (BSV, % CV) | 22.4% | shrinkage 3.2% |  |

BSV, between-subject variability; %RSE, relative standard error.

**Table S3. Population pharmacokinetic model comparison: linear 2-compartment versus 2-compartment with Michaelis-Menten elimination.** Models were fitted using FOCEI in nlmixr2est to the pooled dataset comprising 104 participants and 816 plasma isoniazid concentrations across both study occasions. The Michaelis-Menten model was selected based on lower AIC and BIC values. ΔAIC = 1571.5 − 1450.5 = +121 for the linear model relative to Michaelis-Menten (the selected model has the lower AIC). Values of |ΔAIC| >10 indicate strong model preference.

| **Model** | **Structure** | **k** | **OFV (-2LL)ᵃ** | **AIC** | **BIC** | **ΔAIC** |
| --- | --- | --- | --- | --- | --- | --- |
| **Linear 2-compartment** | First-order elimination with two-compartment disposition | 9 | 112.6 | 1571.5 | 1613.5 | +121 |
| **2-compartment + Michaelis-Menten (selected)** | Saturable elimination (V_max_, K_m_) with two-compartment disposition | 10 | -10.4 | 1450.5 | 1498.3 | Reference |

OFV, objective function value (-2 log-likelihood); k, number of estimated parameters; AIC, Akaike information criterion; BIC, Bayesian information criterion; FOCEI, first-order conditional estimation with interaction. ᵃThe OFV column reports the nlmixr2 objective function value directly and is offset from the AIC/BIC columns by a constant of 1440.9 (the normalisation constant from the -2LL computation).

Table S4. Within-subject paired comparison of INH exposure (Genotype-guided vs Flat 900 mg). Wilcoxon signed-rank test on paired within-subject differences between Day 7 (genotype-guided) and Day 14 (flat 900 mg) values. The final two rows report the McNemar chi-square test (continuity-corrected) on the binary supratherapeutic endpoint C₂₄ > 0.15 µg/mL, the threshold above which weekly 3HP exposure exceeds the concentration previously associated with systemic drug reactions during 3HP. Cell entries for the final two rows are n (%).

| **Parameter** | **NAT2 phenotype** | **N pairs** | **Genotype-guided, median (IQR) or n (%)** | **Flat 900 mg, median (IQR) or n (%)** | **p-value** |
| --- | --- | --- | --- | --- | --- |
| **AUC₀–₂₄ (mg-h/L)** | Slow | 42 | 18.88 (11.88-24.80) | 70.74 (45.26-95.18) | 2.42e-08 |
| **AUC₀–₂₄ (mg-h/L)** | Intermediate | 45 | 32.99 (25.54-45.95) | 39.24 (23.69-50.72) | 0.248 |
| **AUC₀–₂₄ (mg-h/L)** | Rapid | 16 | 42.35 (29.01-63.75) | 20.21 (11.57-28.75) | 6.1e-05 |
| **AUC₀–₂₄ (mg-h/L)** | All | 103 | 27.19 (18.85-41.34) | 43.18 (27.35-70.99) | 1.55e-05 |
| **Cₘₐₓ (µg/mL)** | Slow | 42 | 2.62 (2.17-4.24) | 9.33 (6.75-13.02) | 2.42e-08 |
| **Cₘₐₓ (µg/mL)** | Intermediate | 45 | 9.51 (6.57-12.15) | 9.80 (6.19-13.00) | 0.291 |
| **Cₘₐₓ (µg/mL)** | Rapid | 16 | 13.26 (7.03-16.44) | 6.54 (3.41-10.57) | 0.00058 |
| **Cₘₐₓ (µg/mL)** | All | 103 | 5.74 (2.77-11.44) | 9.24 (6.20-12.01) | 0.000368 |
| **C₂₄** **(µg/mL)** | Slow | 42 | 0.05 (0.03-0.07) | 0.20 (0.11-0.33) | 3.03e-08 |
| **C₂₄** **(µg/mL)** | Intermediate | 44 | 0.02 (0.01-0.03) | 0.01 (0.01-0.02) | 0.0348 |
| **C₂₄ (µg/mL)** | Rapid | 16 | 0.02 (0.02-0.03) | 0.01 (0.01-0.01) | 0.0012 |
| **C₂₄ (µg/mL)** | All | 102 | 0.02 (0.01-0.05) | 0.02 (0.01-0.19) | 0.000169 |
| **C₂₄ > 0.15 µg/mL, n (%) [McNemar]** | Slow | 42 | 1 (2.4%) | 27 (64.3%) | 9.44e-07 |
| **C₂₄ > 0.15 µg/mL, n (%) [McNemar]** | All | 102 | 5 (4.9%) | 30 (29.4%) | 8.32e-06 |

IQR, interquartile range. Paired analyses include participants with complete pharmacokinetic sampling on both the genotype-guided (Day 7) and flat 900 mg (Day 14) occasions. One slow-acetylator participant completed intensive PK sampling on the genotype-guided occasion only; the flat 900 mg occasion data are absent for this participant, and sample was therefore excluded from all paired analyses (paired slow n = 42; paired AUC₀–₂₄/Cₘₐₓ total n = 103; paired C₂₄ total n = 102). One intermediate-acetylator participant is missing the 24-hour post-dose sample on the flat 900 mg occasion; this participant is included in paired AUC₀–₂₄ and Cₘₐₓ analyses (paired intermediate n = 45) but excluded from paired C₂₄ analysis (paired intermediate n = 44). The McNemar chi-square test (continuity-corrected) for C₂₄ > 0.15 µg/mL uses paired C₂₄ observations (slow n = 42, intermediate n = 44, rapid n = 16; total n = 102).

**Table S5. Sensitivity analysis of per-subject isoniazid exposures derived by non-compartmental analysis (NCA, primary) versus the Michaelis-Menten popPK model (sensitivity).** NCA values are linear-trapezoidal integrals of observed plasma isoniazid concentrations at 1, 2, 8, and 24 hours post-dose. Model-derived values are integrals of dense (1-min grid) individual concentration profiles simulated from the final two-compartment Michaelis-Menten popPK model (MM-IPRED) using each participant’s empirical-Bayes individual parameter estimates (V_max_, K_m_, V_2_, Q, V_3_, K_a_). Reported as median (interquartile range). Note: model-derived exposures depend only on each subject’s individual parameters and dose, with no inter-occasion variability in the structural model.

| **Phenotype** | **Dosing** | **Method** | **AUC₀–₂₄ (mg·h/L)** | **Cₘₐₓ (µg/mL)** | **C₂₄ (µg/mL)** | **% C₂₄ > 0.15 µg/mL** |
| --- | --- | --- | --- | --- | --- | --- |
| **Slow** | Genotype-guided (300 mg) | NCA (primary) | 18.88 (11.88–24.80) | 2.62 (2.17–4.24) | 0.05 (0.03–0.07) | 2.4% |
| **Slow** | Genotype-guided (300 mg) | MM-IPRED | 17.10 (13.62–19.55) | 3.29 (2.80–3.64) | 0.04 (0.02–0.06) | 2.4% |
| **Slow** | Flat 900 mg | NCA (primary) | 70.74 (45.26–95.18) | 9.33 (6.75–13.02) | 0.20 (0.11–0.33) | 64.3% |
| **Slow** | Flat 900 mg | MM-IPRED | 68.93 (55.08–78.06) | 10.13 (8.56–11.20) | 0.22 (0.11–0.32) | 66.7% |
| **Intermediate** | Genotype-guided (900 mg) | NCA (primary) | 32.99 (25.54–45.95) | 9.51 (6.57–12.15) | 0.02 (0.01–0.03) | 8.9% |
| **Intermediate** | Genotype-guided (900 mg) | MM-IPRED | 35.00 (28.89–41.78) | 9.50 (7.84–10.71) | 0.02 (0.01–0.03) | 4.4% |
| **Intermediate** | Flat 900 mg | NCA (primary) | 39.24 (23.69–50.72) | 9.80 (6.19–13.00) | 0.01 (0.01–0.02) | 6.7% |
| **Intermediate** | Flat 900 mg | MM-IPRED | 35.00 (28.89–41.78) | 9.50 (7.84–10.71) | 0.02 (0.01–0.03) | 4.4% |
| **Rapid** | Genotype-guided (1,500 mg) | NCA (primary) | 42.35 (29.01–63.75) | 13.26 (7.03–16.44) | 0.02 (0.02–0.03) | 0.0% |
| **Rapid** | Genotype-guided (1,500 mg) | MM-IPRED | 49.24 (44.64–56.94) | 15.09 (12.94–17.10) | 0.02 (0.02–0.02) | 0.0% |
| **Rapid** | Flat 900 mg | NCA (primary) | 20.21 (11.57–28.75) | 6.54 (3.41–10.57) | 0.01 (0.01–0.01) | 0.0% |
| **Rapid** | Flat 900 mg | MM-IPRED | 23.51 (21.65–27.14) | 8.78 (7.55–9.92) | 0.01 (0.01–0.01) | 0.0% |

NCA, non-compartmental analysis; MM-IPRED, Michaelis–Menten model individual predicted concentrations; IQR, interquartile range. Paired sample sizes follow the same structure as Table S4: paired AUC₀–₂₄ and Cₘₐₓ: slow n = 42, intermediate n = 45, rapid n = 16 (total n = 103); paired C₂₄: slow n = 42, intermediate n = 44, rapid n = 16 (total n = 102). See Table S4 footnote for details of participant exclusions.
